# Virus strain of a mild COVID-19 patient in Hangzhou represents a new trend in SARS-CoV-2 evolution related to Furin cleavage site

**DOI:** 10.1101/2020.03.10.20033944

**Authors:** Xi Jin, Kangli Xu, Penglei Jiang, Jiangshan Lian, Shaorui Hao, Hangping Yao, Hongyu Jia, Yimin Zhang, Lin Zheng, Nuoheng zheng, Dong Chen, Jinmei Yao, Jianhua Hu, Jianguo Gao, Liang Wen, Jian Shen, Yue Ren, Guodong Yu, Xiaoyan Wang, Yingfeng Lu, Xiaopeng Yu, Liang Yu, Dairong Xiang, Nanping Wu, Xiangyun Lu, Linfang Cheng, Fumin Liu, Haibo Wu, Changzhong Jin, Xiaofeng Yang, Pengxu Qian, Yunqing Qiu, Jifang Sheng, Tingbo Liang, Lanjuan Li, Yida Yang

## Abstract

The outbreak of COVID-19 become enormous threat to human beings, showing unclear virus mutation during dissemination. We found, in our 788 confirmed COVID-19 patients, the decreased rate of severe/critical type, increased liver/kidney damage and prolonged period of nuclear acid positivity, when compared with Wuhan. To investigate underlining mechanisms, we isolated one strain of SARS-CoV-2 (ZJ01) in mild COVID-19 patient and found the existence of 35 specific gene mutation by gene alignment. Further phylogenetic analysis and RSCU heat map results suggested that ZJ01 may be a potential evolutionary branch of SARS-CoV-2. We classified 54 strains of viruses worldwide (C/T type) based on the base (C or T) at positions 8824 and 28247. ZJ01 has both T at those sites, becoming the only TT type currently identified in the world. The prediction of Furin cleavage site (FCS) and the sequence alignment of virus family indicated that FCS may be an important site of coronavirus evolution. ZJ01 had mutations near FCS (F1-2), which caused changes in the structure and electrostatic distribution of S protein surface, further affecting the binding capacity of Furin. Single cell sequencing and ACE2-Furin co-expression results confirmed that Furin level was higher in the whole body, especially in glands, liver, kidney and colon while FCS may help SARS-CoV-2 infect these organs. The evolutionary pattern of SARS-CoV-2 towards FCS formation may result in its clinical symptom becoming closer to HKU-1 and OC43 (the source of FCS sequence-PRRA) caused influenza, further showing potential in differentiating into mild COVID-19 subtypes.

## Introduction

The outbreak of a novel coronavirus (SARS-CoV-2) and its infected Disease (COVID-19) in Wuhan since the end of 2019 and quickly spreading over the whole country, has put severe threat on public health and economic retardation to China^1, 2^. Through quick response and drastic measures even including quarantining Wuhan city on Jan 23, 2020, the spreading of SARS-CoV-2 was effectively controlled. Nevertheless, its ensuing global sporadic appearance^3, 4^ and quick dissemination in Japan, South Korea, Iran and Italy raised world alert of SARS-CoV-2 pandemics^5, 6, 7^. Therefore, it is urgent to continuingly illustrate the clinical and virological characteristics of SARS-CoV-2 during its dissemination.

An important and common feature of virus is that its increased transmissibility usually accompanies with decreased virulence, which also holds true for SARS-CoV-2 and is reflected in the disease trajectory. On one hand, COVID-19 was more severe in Wuhan at its early stage, reaching approximately 32% of severe/critical types and 11% of fatality^8, 9^. In contrast, data out of Wuhan showed more mild type of COVID-19, as presented in Zhejiang province^10^ and nationwide^11^. On the other hand, its transmissibility was increased from varied basic reproductive number (R0) of 2.2^12^ and 2.68^6^ based on Wuhan data to that of 3.77^13^ in national level. Furthermore, the observation of similar viral load between COVID-19 patients with and without symptoms revealed its capacity of occult transmission^14^.

It is well acknowledged that change in the epidemiological and clinical features of COVID-19 roots from virological change of SARS-CoV-2, in which its spike (S) surface envelope protein plays an important role^15^. Generally, its surface unit (S1) is responsible for host entry by binding to cell receptor while its transmembrane unit (S2) takes charge of the fusion of viral and cellular membranes^16^. Therefore, it is of great value to focus on the sequence mutation and conformation change in S protein for SARS-CoV-2 evolution in a finely established model, aiming to explain the related change of COVID-19.

In this study, we identified 9.9% severe/critical type in 788 confirmed COVID-19 patients of Zhejiang province and median 11 days of positive nuclear acid in 104 patients from our hospital, showing the tendency of COVID-19 progression towards mild but more infective direction. Based on these clinical findings, we performed in-depth bioinformatics analysis by comparing the virological features of previously reported 52 strains of SARS-CoV-2, including Bat coronavirus, SARS-CoV and SARS-CoV-2 in Wuhan and ZJ01 (SARS-CoV-2 we firstly reported in one patient with mild COVID-19 in Zhejiang province). We demonstarated that the specific mutation of ZJ01 may represent a *de novo* evolution trend of SARS-CoV-2, which might be related with Furin. Besides, the establishment of a novel SARS-CoV-2 categorization system may facilitate our understanding of virus evolution and its influence on disease severity and progression.

## Materials and Methods

### Data sources and ethics

A retrospective study investigating the epidemiological, clinical and virological features of COVID-19 in designated hospitals of Zhejiang province between Jan 17 and Feb 7, 2020 was performed, followed by calculating the period of positive nuclear acid of COVID-19 in our hospital. All patients were diagnosed as COVID-19 according to WHO interim guidance^17^. and the preliminary data were reported to the authority of Zhejiang province. The study was approved by the Clinical Research Ethics Committee of the First Affiliated Hospital, College of Medicine, Zhejiang University (NO. IIT20200005C). Written informed consent was waived by the ethics commission of the designated hospital, as this study was carried out for emerging infectious disease and to be part of a continuing public health outbreak investigation under national authorization. The subtypes of COVID-19 were categorized into mild, severe and critical, as recently reported^11^. The period of positive nuclear acid is defined as the date of confirmed nuclear acid positivity minus the date of confirmed nuclear acid negativity.

### Procedure and virus strain collection

SARS-CoV-2 was confirmed from samples of throat-swab and sputum in our hospital and local center for disease control and prevention (CDC) of Zhejiang province, by real-time RT-PCR^8^. All patients received chest x-rays or CT scan on admission, with excluding other respiratory viruses such as influenza A (H1N1, H3N2, H7N9), influenza B, respiratory syncytial virus, SARS-CoV and MERS-CoV. The epidemiological, anthropometrical, clinical and laboratory data were collected on admission, with specific attention paid to the period between symptom onset and outpatient visiting/PCR confirmation/hospital admission. One strain of SARS-CoV-2 was successfully isolated from the sputum of COVID-19 patient with mild type in our hospital, followed by whole genome sequencing with previously reported method^18^.

### Sequence data collection and alignment of SARS-CoV-2

Current available coronavirus sequences (n=85) were collectively achieved from NCBI viral genome database (https://www.ncbi.nlm.nih.gov/, n=65), Genome Warehouse (https://bigd.big.ac.cn/gwh/, n=12), CNGBdb (https://db.cngb.org/, n=3) and NMDC (http://nmdc.cn/#/coronavirus, n=6). Sequence of ZJ01 (BataCov/Zhejiang/ZJ01/2019) was provided according to our sequencing result. The 52 sequences of SARS-CoV-2 were collected from China (n = 30), Japan (n = 5), Nepal (n = 1), South Korea (n = 1), Australia (n =1), Finland (n=1) and United States of America (n = 13). The collection period was between December 26, 2019 and February 5, 2020. The Furin protein sequence was downloaded from NCBI. Multiple sequence alignment of all coronavirus genomes was performed by using MEGA v7.0.26.

### Phylogenetic and Relative synonymous codon usage analysis

Phylogenetic analysis was performed on a total of 80 coronavirus strains, covering 6 species (human, bat, mink, camel, rat and pig). The source of SARS-CoV-2 comes from 17 cities in 7 countries, with periods between virus outbreak and dissemination (2019.12.23-2020.2.5). The evolutionary history was constructed based on the S protein of coronavirus by Neighbor-Joining method. The bootstrap consensus tree inferred from 2000 replicates was taken to represent the evolutionary history of the taxa analyzed. Branches corresponding to partitions reproduced in less than 30% bootstrap replicates were collapsed. The evolutionary distances were computed using the Kimura 2-parameter method and were in the units of the number of base substitutions per site. Evolutionary analyses were conducted in MEGA7 v7.0.26. Relative Synonmous Codon Usage (RSCU) analysis was applied to compare the differences between 49 strains of SARS-CoV-2 and Homo, with further heat map drawing by MeV 4.9.0. All available coding sequences (minimum>28 kbp) were calculated with Codon W1.4.2.16, followed by inter-relationship calculation based on Euclidean distance method.

### Mutation site analysis and FCS prediction

SimPlot v.3.5.1.<u>15</u> was used to analyze the potential genetic recombination. The visualization of the mutation site between RATG13 and ZJ01 was performed by Multalin (http://multalin.toulouse.inra.fr/multalin/multalin.html). Multiple sequence alignment was applied by using the Muscle (codons) function of MEAG v7.0.26. Genetic mutation sites were analyzed by DNAMAN v9.0.1.116. The visualization of the functional domain distribution of SARS-CoV-2 and S proteins was plotted by IBS v1.0.3. The FCS prediction was carried out in ProP 1.0 Server (http://www.cbs.dtu.dk/services/ProP/)^19^ and presented as Furin score (range 0-1), where the closer to 1, the higher possibility of having FCS.

### Homology modeling and APBS analysis

Target protein was downloaded from NCBI (https://www.ncbi.nlm.nih.gov/protein/1791269092) and corresponding homology models were predicted by SWWISS-MODEL(https://swwassmodel.expasy.org/). Protein sequences alignment and Adaptive Poisson-Boltzmann Solve (APBS) analysis were performed by PyMOL v2.3.3 on Intel i7 9700F processor. APBS was calculated and evaluated by PyMOL v2.3.3, as previously reported^20^.

### Single cell transcriptome data analysis

The raw counts or processed data were download from Tissue Stability Cell Atlas (https://www.tissuestabilitycellatlas.org/), Gene Expression Omnibus (GEO; https://www.ncbi.nlm.nih.gov/). Lung, colon and liver data were respectively obtained from Tissue Stability Cell Atlas^21^, GSE116222^22^ and HCA^23^, including samples of 5 lung, 3 colonic epithelium and 5 hepatic tissues from healthy volunteers and organ donors. Lung and liver data had been processed before downloading, and directly used for data analysis and visualization. For liver data, cells with less than 100 expressed genes and 1500 UMI counts and higher than 50% mitochondrial genome transcript were removed. Genes expressed in less than three cells were also removed.

Normalization and principal component analysis were performed in the R package Seurat^24^, with different datasets based data processing methods. For liver, the first 40 principal components resulted in principal component analysis were used to perform cell clustering and non-linear dimensionality reduction (Uniform Manifold Approximation and Projection, UMAP). For colon, R package Harmony^25^ was used to remove batch effects with default settings. Then we used first 40 components resulted in harmony to perform cell clustering and non-linear dimensionality reduction as same as liver data. Depending on the expression level of cell markers provided in the original article corresponding to the scRNA-seq datasets, we further estimate which cell types the cell clusters belong to. Annotated clusters were then visualized using UMAP plots with “DimPlot” function in Seurat. Normalized gene expression levels were presented in the UMAP and violin plots by R package ggplot2^26^.

## Results

### Epidemiological and clinical characteristics of 788 enrolled COVID-19 patients and ZJ01 patient

As shown in Table 1, 51.65% of the 788 enrolled patients were male, with low rate of smoking (6.85%). The top three co-existing conditions were hypertension (15.99%), diabetes (7.23%) and chronic liver disease (3.93%). The median period from illness onset to outpatient visiting, PCR confirmation and hospital admission were 2, 4 and 3 days, respectively. The most common symptoms were fever (80.71%) and cough (64.21%) while the highest rate of CT/X-ray manifestation was bilateral pneumonia (37.56%). The rates of mild, sever and critical types of COVID-19 were 90.1%, 7.74% and 2.16%, respectively. The ZJ01 patient is male, 30y, and had neither smoking history nor any co-existing condition. He visited outpatient clinics 1 day after symptom onset and was admitted in hospital confirming with COVID-19 in the same day by PCR test. He had no exposure history to Wuhan and none of his family members was reported to be virus positive till now. Consistent with other COVID-19 patients, he had the symptoms of fever, cough and sputum production, with CT image showing bilateral pneumonia. This patient was categorized into mild type of COVPD-19, with normal blood routine test and examination of inflammation marker (CRP and PCT). The higher level of ALT and serum creatinine in this patient indicated potential liver and kidney injury. Unusually, the period of continuing positive nuclear acid was 24 days, longer than most patients reported from Wuhan^27^.

**Table 1.** Demographic, epidemiologic and clinical characteristics of 788 COVID-19 patients and detailed information on ZJ01 patient.

| **Characteristics** | **All patients**<br>**(% or 1<sup>st</sup> -3<sup>th</sup> quarter)** | **ZJ01**<br>**Patient** |
| --- | --- | --- |
| **Age** | 45.83±14.88 | 30 |
| **Male Sex** | 407(51.65%) | male |
| **Current Smoker** | 54(6.85%) | no |
| **Coexisting Condition** |  | no |
| Hypertension | 126(15.99%) |  |
| Diabetes | 57(7.23%) |  |
| Chronic liver disease | 31(3.93%) |  |
| Chronic renal disease | 7(0.89%) |  |
| Heart disease | 11(1.40%) |  |
| COPD | 3(0.38%) |  |
| **Date of symptom onset to** |  |  |
| Outpatient visiting | 2(1-4) | 1 |
| PCR confirmation | 4(2-7) | 1 |
| Hospital admission | 3(1-6) | 1 |
| **Severity on admission** |  |  |
| Mild | 710(90.1%) | yes |
| Severe | 61(7.74%) |  |
| Critical | 17(2.16%) |  |
| **Fever(yes/no)** | 636/152(80.71%) | yes |
| **Cough** | 506/282(64.21%) | yes |
| **Sputum production** | 265/523(33.63%) | yes |
| **Sore throat** | 111/677(14.09%) | no |
| **Muscle ache** | 91/697(11.55%) | no |
| **GI symptoms** | 78/700(9.89%) | no |
| **Headache** | 66/722(8.38%) | no |
| **Laboratory test** |  |  |
| Leucocytes (×10<sup>9</sup> per L) | 4.8(3.8-6.0) | 4.2 |
| Neutrophils(×10<sup>9</sup> per L) | 2.96(2.22-4.0) | 2.5 |
| Lymphocytes(×10<sup>9</sup> per L) | 1.19(0.9-1.56) | 0.9 |
| INR(normal range 0.85-1.15) | 1.02(0.97-1.09) | 1.01 |
| ALT(U/L) | 21.09(15.0-33.0) | 57 |
| AST(U/L) | 25.0(19.6-33.0) | 34 |
| Serum creatinine(μmol/L) | 66.0(55.2-78.0) | 108 |
| Creatine kinase(U/L) | 69.0(47.0-107.0) | 75 |
| C-reactive protein(mg/L;) | 8.0(2.4-21.2) | 6.5 |
| Procalcitonin, (ng/mL) | 0.05(0.03-0.07) | 0.07 |
| **Chest x-ray/CT findings(yes/no)** |  |  |
| Normal | 87/701(11.04%) |  |
| Unilateral pneumonia | 164/624(20.81%) |  |
| Bilateral pneumonia | 296/492(37.56%) | Yes |
| Multiple mottling and ground-glass opacity | 235/553(29.82%) |  |

### Phylogenetic and RSCU analyses revealed potential evolutionary divergence in SARS-CoV-2 and the unique feature of ZJ01

Phylogenetic analysis suggested that SARS, RATG13 and SARS-CoV-2 exhibited remarkable evolutionary divergence, with potential evolutionary branches within SARS-CoV-2 (Fig 1A). For instance, there existed minor evolutionary divergence between WIV02 (2019-12-31)/WH19008 (2019-12-30) and MT0270641 (2020-1-29)/ZJ01(2020-1-23) that were respectively collected in the early and widely spread stages of the epidemic, indicating the potential of forming evolutionary branches during dissemination. The relative synonymous codon usage (RSCU) analysis showed that there were various differences among 8 strains (MN938384, CNA0007334, WIV06, ZJ01, NMDC60013002-05, CNA0007332, MN988668, WIV07) and other members of SARS-CoV-2 family (Fig 1B), where MN938384, CNA0007334, WIV06 and ZJ01 were the closest to human RSCU. Among aforementioned 8 strains which were collected in Wuhan, Guangdong and Hangzhou, six were collected at the early stage of COVID-19 (2019-12-26 to 2020 −1-2). The collection time of MN938384 was no later than 2020-1-14 (virus submission time), while ZJ01 was collected on 2020-1-23.

**Figure 1.**
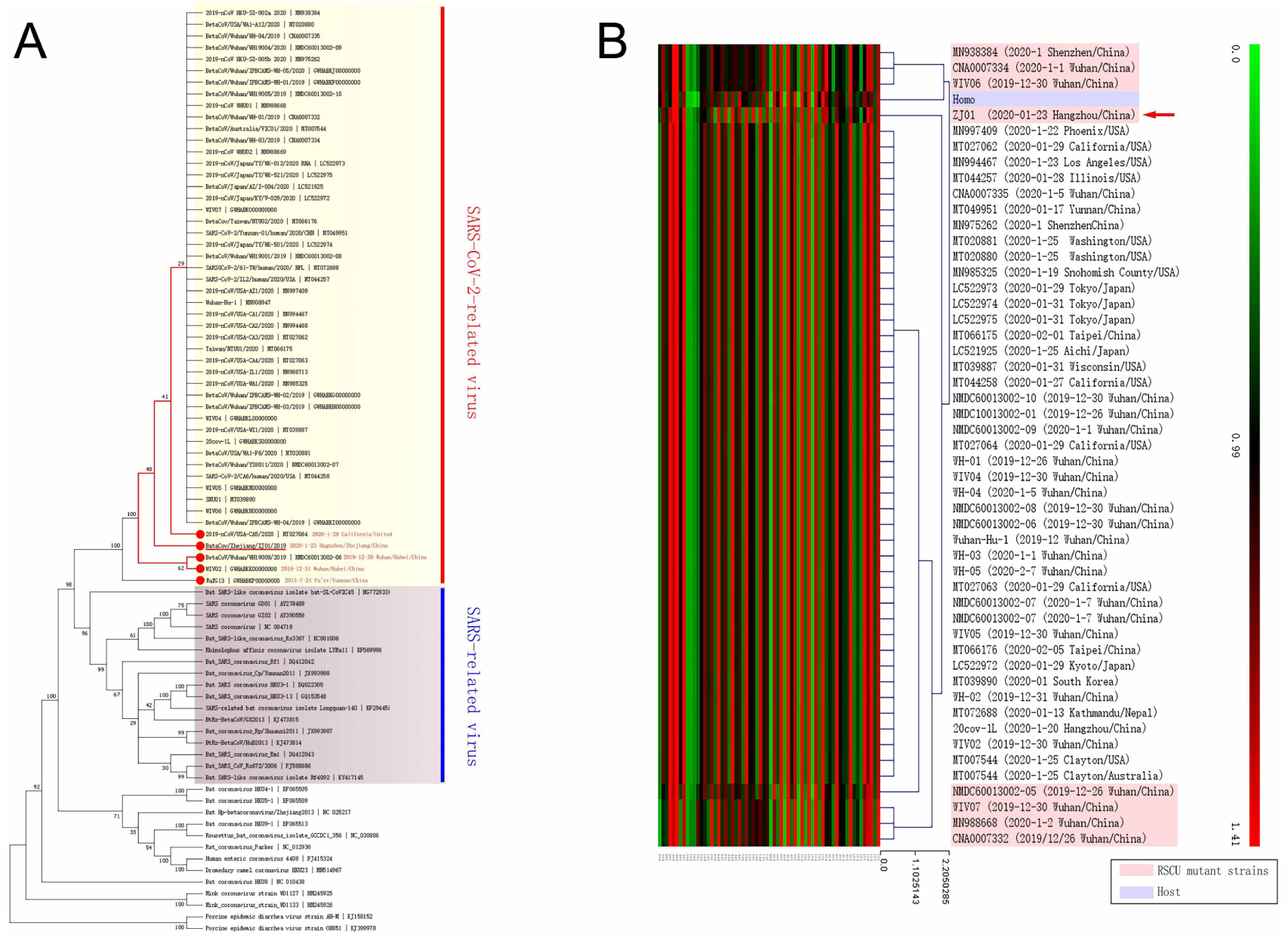
The existence of potential evolutionary divergence in SARS-CoV-2 family. (A), Phylogenetic analysis indicates that SARS and SARS-CoV-2 are totally separated branches in evolution while ZJ01 may be a representative of a potential evolutionary subtype of the SARS-CoV-2 family. (B), RSCU heatmap indicates that 8 strains of SARS-CoV-2 family are more close to human RSCU, differing from other strains. The RSCU of ZJ01 might be one representative of potential branches.

### Sequence alignment analysis identified ZJ01 as a novel viral pattern of SARS-CoV-2

The whole sequence of ZJ01 was deposited in Appendix. Sequence alignment analysis suggested that ZJ01 had 38 mutation sites compared with other SARS-CoV-2 family members (Fig 2A), in which 35 mutations were unique to ZJ01, including 7 deletions, 4 insertions and 24 substitutions. For the rest 3 mutation sites, NO.20 was caused by a sequencing error while NO.14 and No. 38 widely existed in SARS-CoV-2 family. Among ZJ01 unique mutations, 10 (NO.22-31) were located on the S protein, including 3 samesense mutation, 2 deletion mutation and 5 missense mutation, which led to amino acid changes of Ser596, Gln613, Glu702, Ala771, Ala1015, Pro1053 and Thr1066. Further similarity analysis indicated that the main difference among various coronaviruses located in the Receptor Binding Domain (RBD) region of S1. Intriguingly, the differences between ZJ01 and other members of SARS-CoV-2 mainly resided in in the S1/S2 and S2 (Fig 2B).

**Figure 2.**
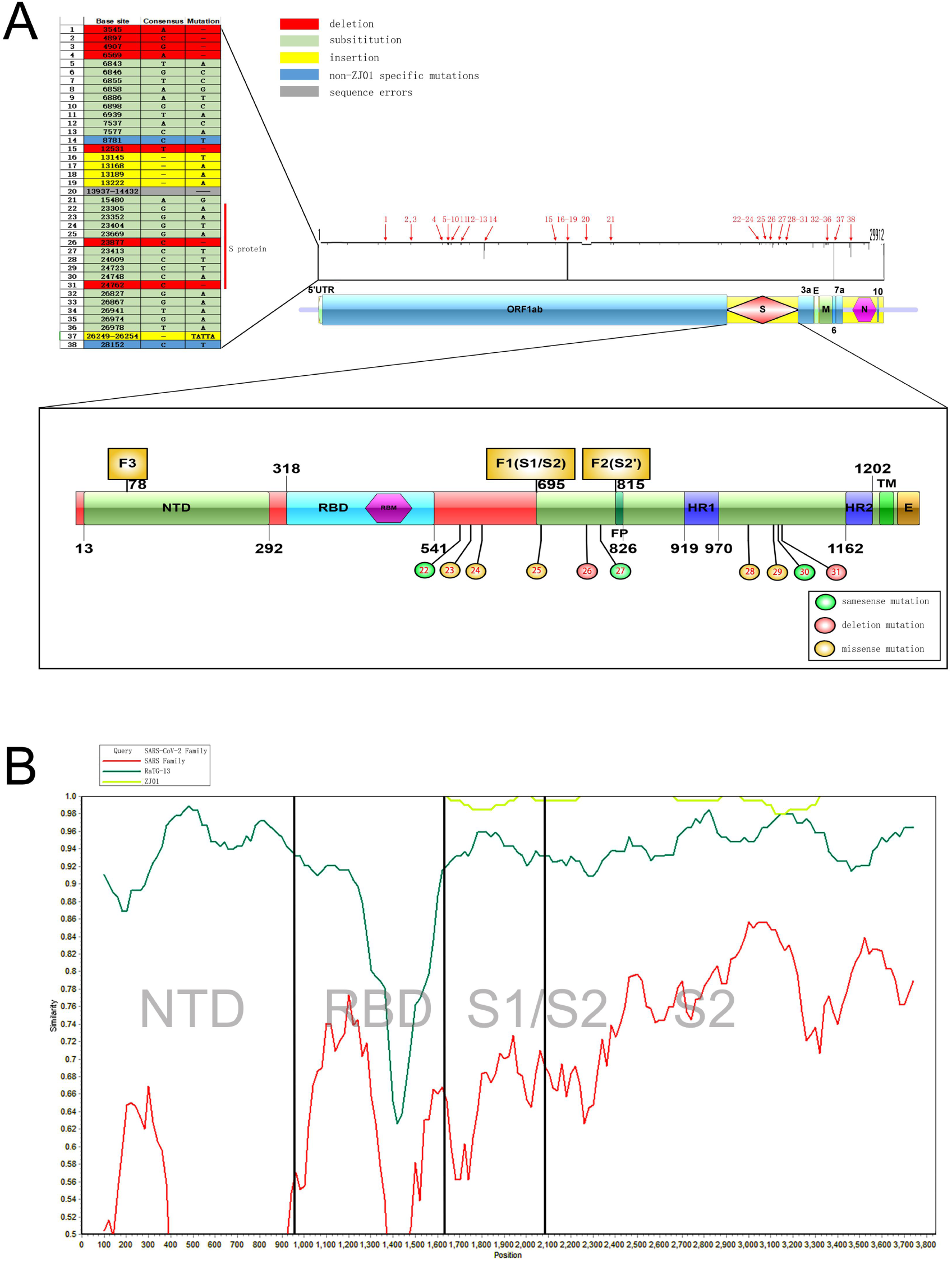
The mutations of ZJ01 when comparing with other strains of SARS-CoV-2 by sequence alignment. (A), ZJ01 has a large number of base mutations far above average while some mutations are unique for ZJ01 and appears across every structural domain. Among them, 10 base mutations appears on the S protein, where 7 cause amino acid changes. (B), Coronavirus mutations between different species are mainly concentrated in the S1 segment, while the mutations of ZJ01 and other members of SARS-CoV-2 are mainly concentrated in the S2 segment.

Further analysis showed the significant association of gene mutation between nucleotide positions 8824 and 28247 in SARS-CoV-2 family (Fig 3A). In detail, when the 8824 was T, its corresponding 28247 was C and vice versa. This pattern was confirmed in 52 out of 54 (96.3%) strains of SARS-CoV-2 and the only two exceptions were from ZJ01 (8824T/28247T) and MN988713 (8824Y/28247Y), where Y meant C or T. Interestingly, Bat coronavirus RATG13 (currently most similar to SARS-CoV-2) and Pangolin coronavirus MP789 (suspicious intermediate host of SARS-CoV-2) both had 8824C/28247T. There were near 20,000 base distance between these two nucleotide positions, and no functional correlation was found, where 8824 was in the middle of ORF1ab and 28247 was nearby N domain. Since mutation could not explain the perfect correlation between the base change of 8824 and 28247 nucleotide positions in 52 strains of SARS-CoV-2, the largest possibility was that certain strain of SARS-CoV-2 carrying 8824C/28247T had a mutation in early stage of dissemination, becoming 8824T/28247C. Those two ancestor strains kept replication during virus spreading and forming their own cladistics (Fig 3B). Though it is still unclear whether the appearance of 8824T/28247C happened in intermediate host stage or human infection stage, we speculated that SARS-CoV-2 kept mutation in human transmission and formed the specific strain of ZJ01 (8824T/28247T).

**Figure 3.**
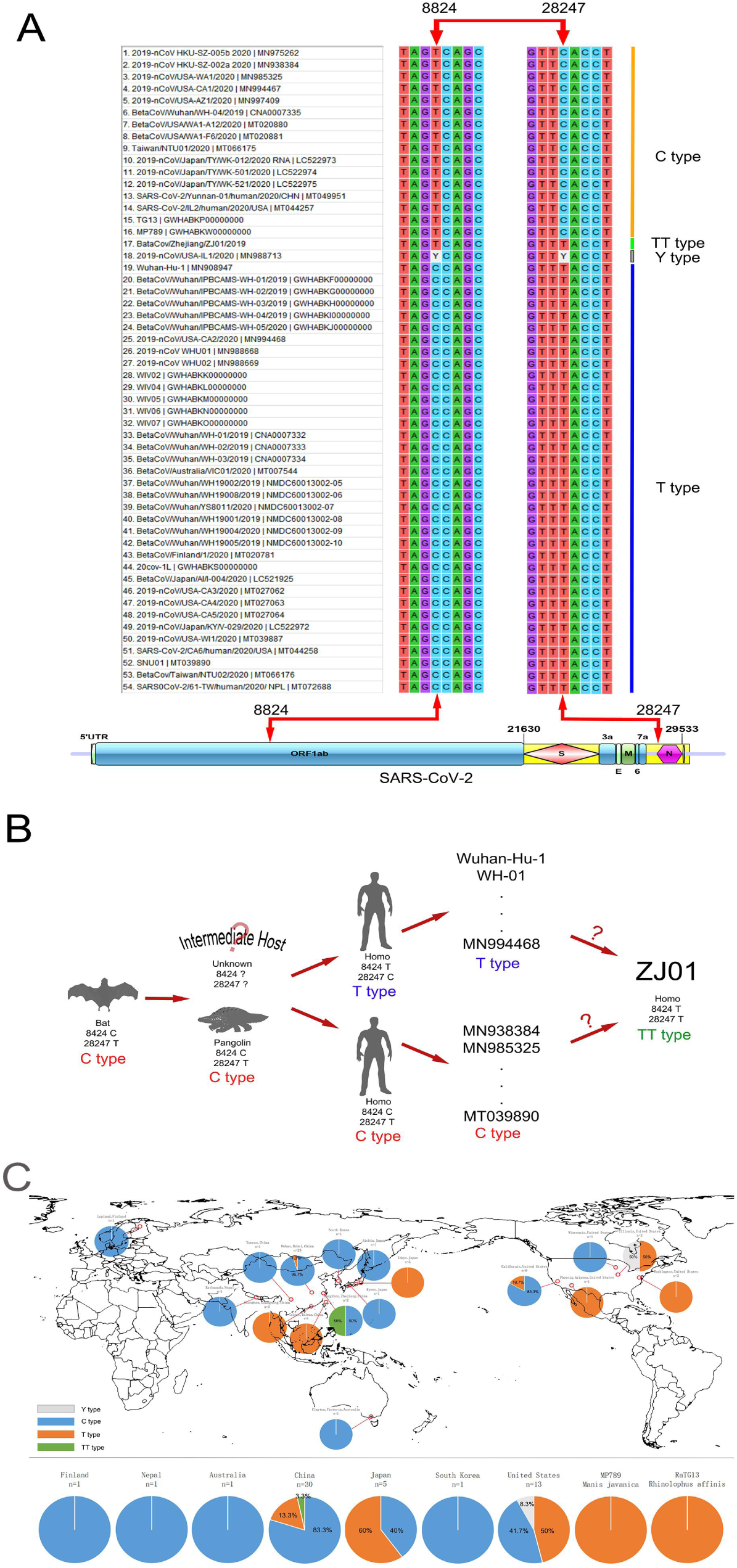
Proposing a C/T categorization system for SARS-CoV-2. (A), SARS-CoV-2 family had base mutation in 8824 and 28247 loci with high correlation of 96.3%. When T appeared in one locus, C would appear in another and vice versa. According to this pattern, we categorized SARS-CoV-2 into type C (68.5%), type T (27.8%), type TT (1.9%) and type Y (1.9%) of available 54 strains. (B), BaTG13 and MP789, the possible ancestor of SARS-CoV-2, was belong to type C. Therefore, the C/T pattern may appear in the intermediate host or early human transmission stage. (C), From the angle of global C/T categorization, China, especially Wuhan, presented with T type while Japan and the USA presented with C type. The difference in the rate of C/T type may help tracing the transmission route of SARS-CoV-2.

We proposed a novel categorization system for SARS-CoV-2, and defined type C for 8824C, type T for 8824T and type TT for ZJ01 as a special case, respectively. According to this system, we further categorized 54 strains of SARS-CoV-2-related virus (Fig 3C). We found 83.3% and 95.7% T type in China (n=30) and Wuhan (n=23); 60% and 100% C type in Japan (n=5) and Tokyo (n=3); 53.8% C type in the USA (n=13), 83.3% T type in California (n=6) and 100% C type in Washington D.C (n=3). Intriguingly, of two cases from Illinois, one was T type and the other was named as Y type for Y in both nucleotide positions 8824 and 28247, indicating the possibility of co-infection with both T and C types. Furthermore, TT type was only reported in Hangzhou. Whether this is an occasional single mutated strain or a novel potential subtype of SARS-CoV-2 warrant more in-depth virological analysis in the future.

### Evolutionary divergence in SARS-CoV-2 strongly correlated with Furin protein

There were 3 potential furin cleavage site (FCS) on S protein, where F1, F2 and F3 separately located in S1/S2, S2 and the N-terminal domain (NTD) of S1 (Fig 4A). Further comparative alignment analysis on GZ02 (SARS viral strain), Wuhan-Hu-1 (the earliest sequenced SARS-CoV-2), RaTG13, HKU9-1 (the potential ancestor of SARS and SARS-CoV-2) and SARS-CoV-2 (HKU-1 and OC43) showed that FCS exhibited a clear sequence and rule in coronavirus evolution (Fig 4B). In detail, there was no FCS in HKU9-1, but one potential FCS in F2 locus of GZ02 (Furin score 0.366) showed effective furin binding capacity^28^. For RaTG13, F2 locus was slightly changed (Furin score 0.333) and a novel FCS was formed in F1 locus (Furin score 0.279). Though the changes in these two sites were inherited in SARS-CoV-2, there still existed huge differences in F1 site between RaTG13 and SARS-CoV-2.

**Figure 4.**
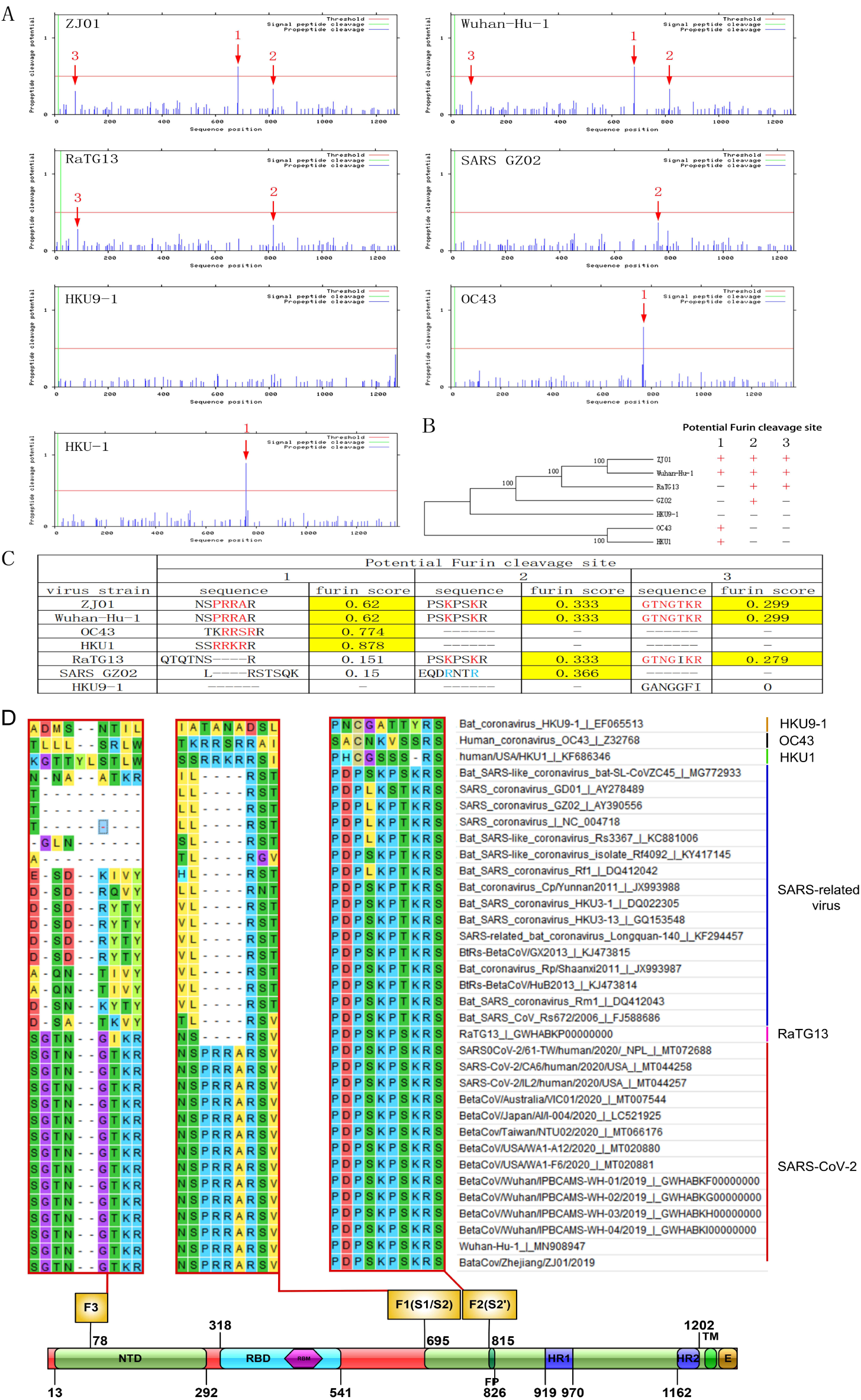
The important role of FCS in SARS-CoV-2 evolution. (A), The number of FCS was varied in different coronavirus. SARS-CoV-2 had F1-3 sites, RaTG13 had F1-2 sites with 96% similarity to SARS-CoV-2, SARS had F2 site, HKU1 and OC43 had F1 site while HKU9-1 had no FCS. (B), the location and number of FCS may play an important role in coronavirus evolution. (C), The sequence of F1 shared high similarity among HKU1, OC43 and SARS-CoV-2, which may be caused by the Furin cleavage site translocation during the evolution of SARS-CoV-2 ancestor and finally resulted its human transmission. (D), F1-3 showed high conservation in different coronavirus, hinting the potential role in virus invasion and replication.

Strikingly, we found an additional PRRA sequence in the F1 site of SARS-CoV-2 when compared with RaTG13, forming a strong and reliable FCS (Furin score 0.62). Though the source of insertion was unknown, the PRRA sequence was common in Avian Influenza virus^29^. We deduced that its source might inherit from HKU1 and OC43, which had effective FCS in F1 site (Furin score 0.878 and 0.744) and respective amino acid of SSRRKRR and TKRRSRR, with high similarity of NSPRRAR in SARS-CoV-2. HKU1 and CO43 could cause human upper respiratory tract infection but the symptom was milder than SARS and SARS-CoV-2. Therefore, we reckoned that the ancestor of SARS-CoV-2 may exchange gene with HKU1 or OC43 to obtain FCS in F1 site during evolution to human transmission (Fig 4C). ZJ01 had Glu702 to Lys702 substitution at the site 18^th^ amino acid behind F1 site, and deletion (Ala771 to -) at the site 37^th^ amino acid ahead of F2 site. These mutations may influence the tertiary and quaternary structure of S protein and finally change the Furin binding capacity. F1-3 sites were conservative in SARS-CoV-2 and SARS (Fig 4D), indicating the importance of mutation in these sites.

### Protein structure analyses implied the mutation in F1 and F2 related area of ZJ01 may influence its binding capacity with Furin protein

Homology modeling clearly revealed the position of F1-3 sites in S protein of SARS-CoV-2 (Fig 5A). Briefly, F1-3 all located on the surface of S protein and protruded outward, exhibiting high potential as substrate binding sites. F1 located on the transition area of S1 and S2 (S1/S2) with obvious outward protrusion, F2 located on the mid-lower position of S2, while F3 located on the top of S1-NTD. Further homology modeling of the S protein of GZ02, RaTG13, Wuhan-Hu-1 and ZJ01 showed big differences in protein structure conformation of F1 locus. From SARS, RaTG13 to SARS-CoV-2, the F1 site showed the tendency of outward protrusion (Fig 5B). Though Wuhan-Hu-1 and ZJ01 shared same amino acid sequence in F1 site, the mutation (Glu702 to Lys702) nearby F1 site of ZJ01 may still change its protein structure conformation and result in further outward extension by 11.6 Å. Furthermore, RaTG13, Wuhan-Hu-1 and ZJ01 owned high degree of consistency in F2 site. The F2 site of GZ02 was deeply buried in the inner place of S protein, that was the biggest difference of SARS-CoV-2 whose F2 site was on the surface of S protein. Finally, RaTG13, Wuhan-Hu-1 and ZJ01 showed high similarity in F3 site that was missing in GZ02.

**Figure 5.**
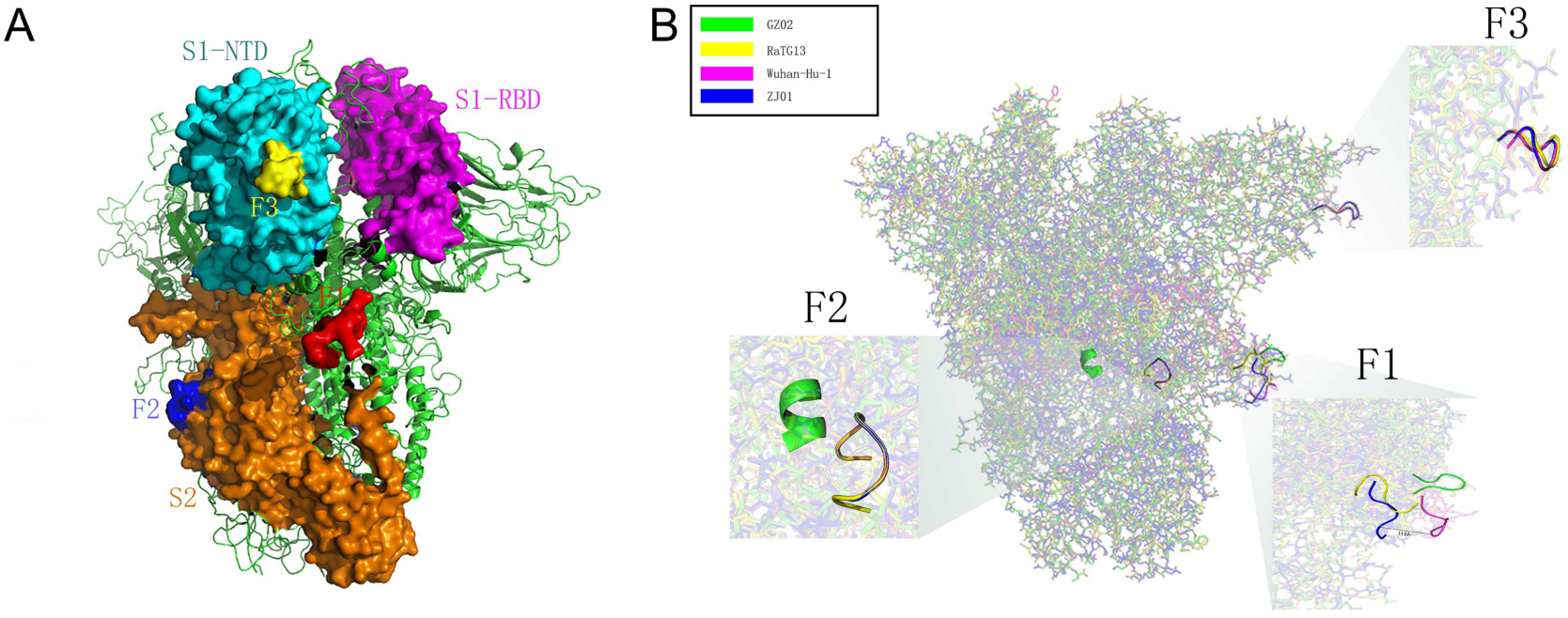
The spatial location and protein structure of potential FCS. (A), The spatial position of F1-3 on S protein. F1 locates at S1/S2, F2 locates at S2 and F3 locates at NTD of S1. (B), There are differences in the tertiary structure of the protein at the F1-3 sites of GZ02, RaTG13, Wuhan-Hu-1, and ZJ01. The difference between ZJ01 and Wuhan-Hu-1 may be caused by the mutation of ZJ01 near FCS.

Adaptive Poisson-Boltzmann Solver (APBS)analysis revealed that furin was a protease with negative charge. Its substrate binding site (191-192, 253-258, 292-295) was covered with a large number of negative charges (Fig 6). The F1 site from SARS-CoV-2 related virus (ZJ01, Wuhan-Hu-1 and RaTG13) were predominantly covered with positive charge while SARS was mixed with negative and positive charge. Compared with Wuhan-Hu-1, F1 site of ZJ01 had more positive charge in its protruding head and more negative charge in its basal part. The F2 site of GZ02 was covered with negative charge while F2 site of Wuhan-Hu-1 and RaTG13 were covered with low level of positive charge. ZJ01 had more positive charge in F2 site than the other strains, probably caused by nearby gene deletion (Ala 771 to -). GZ02 had many negative charges in F3 site while few negative charges were identified in SARS-CoV-2 related virus. Therefore, we speculated that, though ZJ01 shared gene similarity with Wuhan-Hu-1, its mutation near FCS changed protein structure conformation and surface electrostatic potential, which further influenced its binding capacity with Furin.

**Figure 6.**
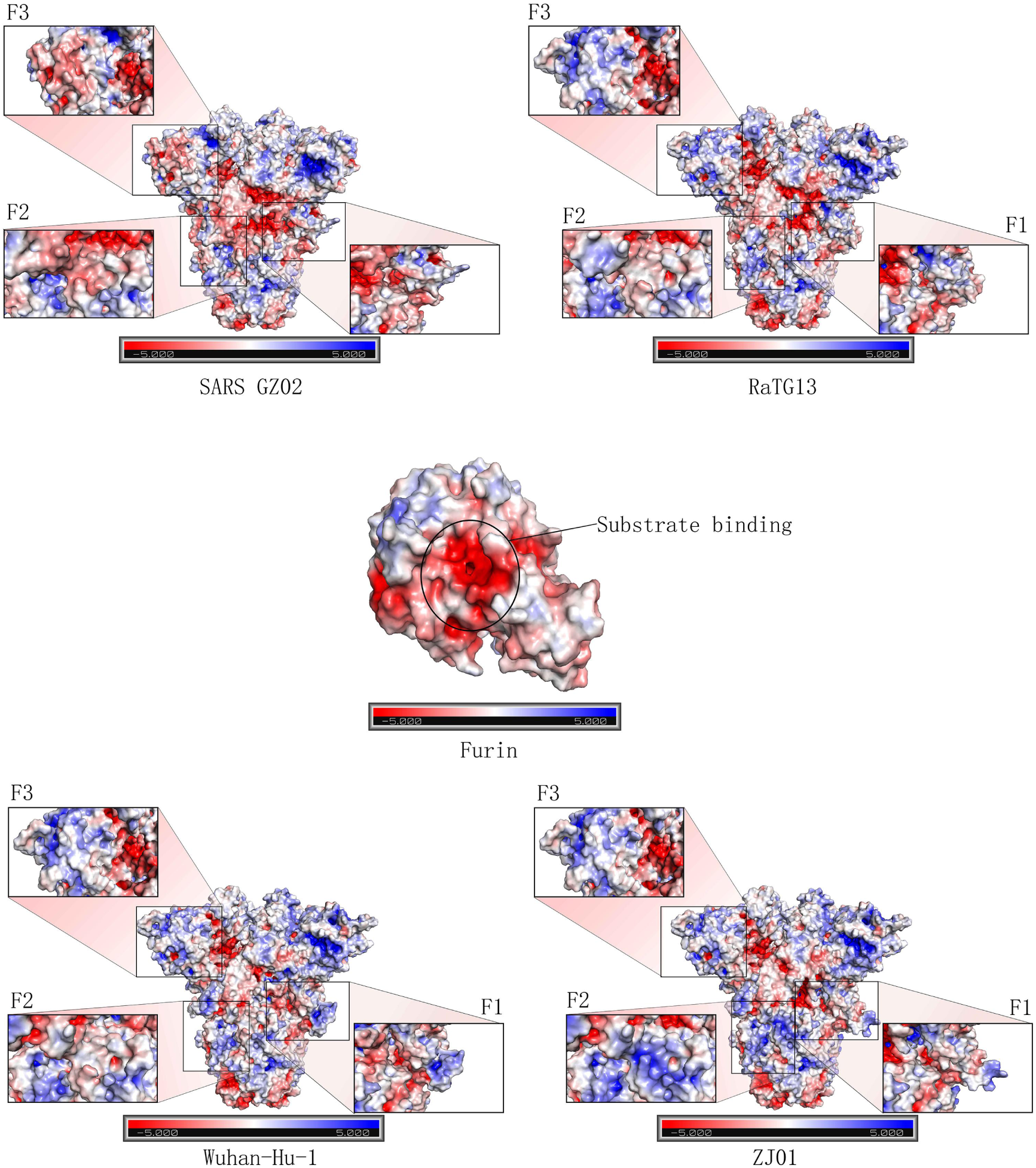
APBS analysis indicates the mutation near Furin cleavage site may cause surface electrostatic potential change of S protein. Furin presents with negative charge, especially its inner substrate-binding pocket. The APBS analysis on the F1-3 sites of GZ02, ZJ01, Wuhan-Hu-1 and RaTG13 reveals the existence of great difference in electrostatic potential, which may be caused by protein structure change in F1-3 and nearby locus. The mutation in F1-2 sites of ZJ01 may cause its differed binding capacity with Furin, when compared with Wuhan-Hu-1.

### Higher expression of Furin than ACE2 in different organs at single cell level

The protein and RNA expression level of ACE2 and Furin in human major tissues were explored in The Human Protein Atlas (https://www.proteinatlas.org/). We found that ACE2 was predominantly expressed in small intestine, duodenum, colon, kidney and testis, while was relatively lowly expressed in lung (Fig. 7A). Furin was expressed in most tissues and organs of the human body, and exhibited highest expression levels of RNA in salivary gland, placenta, liver, pancreas and bone marrow (Fig. 7B). Of note, Furin had extremely low protein expression level in lung compared with other tissues.

**Figure 7.**
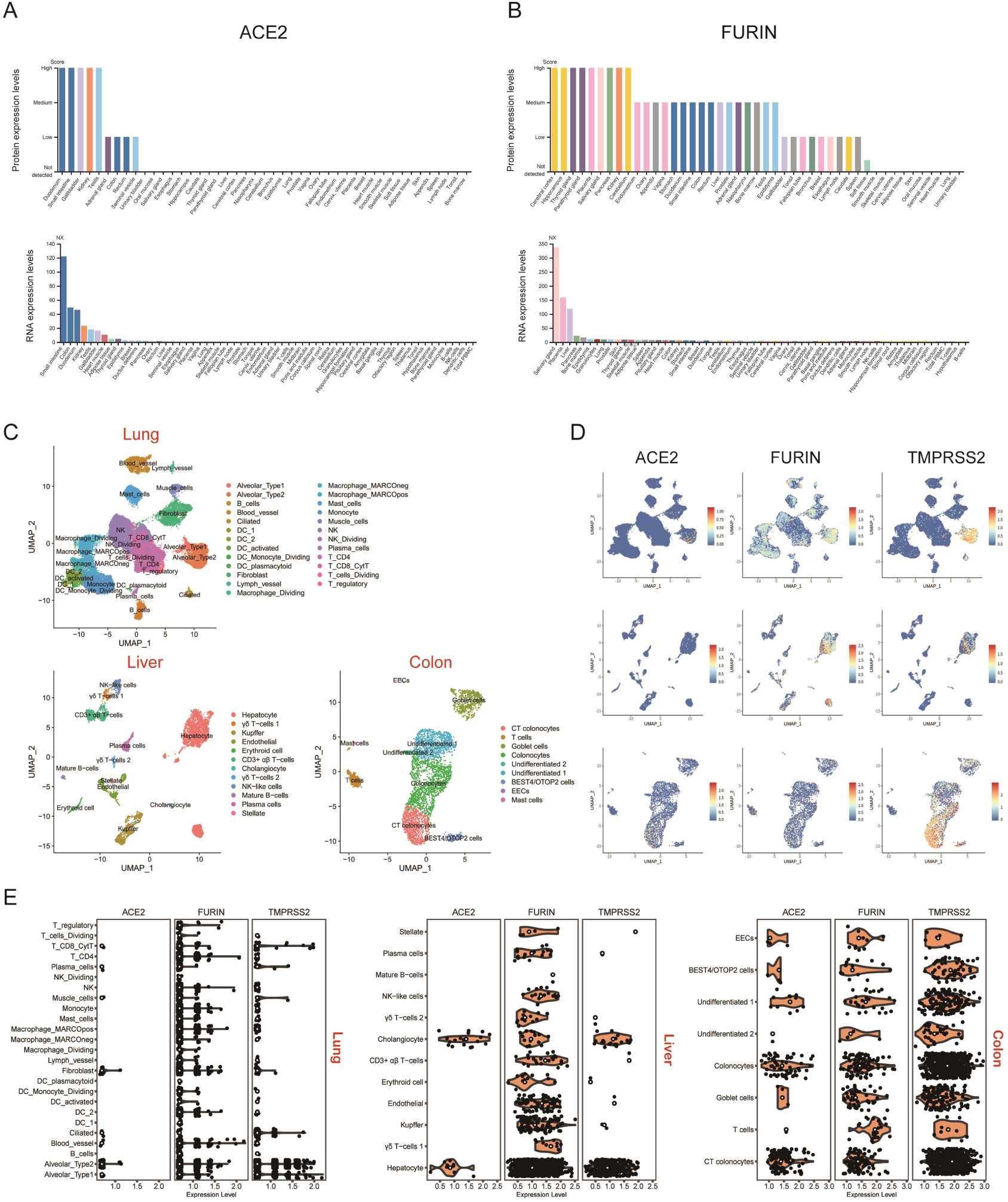
The expression profiles of ACE2 and FURIN in multiple human organs. The protein and RNA expression levels of (A) ACE2 and (B) FURIN in human major tissues. NX: Normalized eXpression. (C), UMAP plots showed cell types identification in lung, liver and colon scRNA-seq data. DC, dendritic cell; CT, crypt top; ILCs, innate lymphoid cells; EECs, enteroendocrine cells; BEST4/OTOP2 cells, cells with high expression of BEST4 and OTOP2. (D), UMAP plots showed ACE2, FURIN and TMPRSS2 specifically expressed cells in lung, liver and colon. (E), Violin plots showed ACE2, FURIN and TMPRSS2 expression levels in different cells types of lung, liver and colon. Black dots represent the level of gene expression in each cell, white circles represent median values.

To further explore the correlation between ACE2 and FURIN expression, we reanalyzed single-cell RNA sequencing (scRNA-seq) data in lung, liver, colon as described in Methods (Figure 7C). Since ACE2 and TMPRSS2 co-expression were reported recently^30^, we also examined TMPRSS2 expression levels in these tissues. In the scRNA-seq datasets, ACE2, FURIN and TMPRSS2 showed higher expression levels in liver or colon than in lung (Fig. 7E). Consistently with previously report^31^, ACE2 was mainly expressed in alveolar type 2 cells in lung (Fig. 7D-E). ACE2 was highly expressed in liver cholangiocytes, liver hepatocytes, colon colonocytes and colon CT colonocytes compared with other liver or colon cell types respectively. This expression pattern was the same as TMPRSS2, but TMPRSS2 had a higher expression level in each cell type. In contrast, FURIN was expressed in all cells types of three tissues, but had little co-expression with ACE2.

## Discussion

The outbreak of COVID-19 has been rapidly spreading over the recent 2 months, causing enormous damage to China. With nationwide dissemination, its epidemiological and clinical features have changed. Accumulating evidences indicate the appearance of several unique characteristics distinct from Wuhan^8, 9, 32^, including higher rate of mild type, lower rate of severe/critical type and mortality and longer period of nuclear acid positivity^10, 11, 27, 33^. Moreover, the increased transmission route of SARS-CoV-2 has been gradually unmasked, from previous recognition of respiratory transmission to through feces^34^ and even tears and conjunctival secretions^35^ All these data indicate the possibility of virus evolution during its spreading, towards the direction of decreased severity but increased transmissibility.

However, according to recently published virus sequencing results^18^, the SARS-CoV-2 family members shared similar gene sequences and few, if there is any, essential changes occurred. How to explain the contrary phenomenon in change of clinical features and virus conservation? We selected one COVID-19 patient with mild type and isolated his virus (ZJ01) for comparative analysis. We found a total of 37 gene mutations, with 35 unique to ZJ01. Further bioinformatics analysis showed the difference between ZJ01 and other strains of SARS-CoV-2 as well as the important roles of Furin. Thereafter, we conclude that SARS-CoV-2 is undergoing evolution towards mild direction with increased furin cleavage sites.

Through analysis on 788 COVID-19 patients in Zhejiang province, we found that there existed mild and severe types of SARS-CoV-2. Though we didn’t currently have evidence to prove whether patients with mild COVID-19 are directly caused by virus mutation or other factors, we indeed found the significant difference between ZJ01 and other members of SARS-CoV-2. ZJ01 had relative high number of 37 mutations and its RSCU was closer to humans than most members of SARS-CoV-2. More importantly, ZJ01 was the only TT type of total 54 strains in our C/T categorization system. Though the sequence of ZJ01 was still close to Wuhan-Hu-1 (the earliest identified SARS-CoV-2) and its mutations were not sufficient to reach the threshold of forming an independent subtype, our evidence indicated ZJ01 may represent a specific evolution direction of SARS-CoV-2.

In this study, we first developed the C/T categorization system of SARS-CoV-2, which revealed the occurrence of possibly inheritable mutation at the very early stage of its evolution and the potential of continuing C/T subtype formation. The TT type of ZJ01 was unique in our system. Though a similar categorization system has been recently proposed^36^, they did not report such TT type in their 120 strains of SARS-CoV-2. Besides, the C/T pattern could also be used to trace the route of virus infection and evolution. For instance, we found 8 strains of T type with 29198T, including RaTG13, MP789, 2019-nCoV_HKU-SZ-005b_2020 and 2019-nCoV_ HKU-SZ-002a_2020 from ShenZhen (China), 2019-nCoV/USA-AZ1/2020 from Arizona/Phoenix (the USA), 2019-nCoV/Japan/TY/WK-501/2020, 2019-nCoV/Japan/TY/WK-012/2020 and 2019-nCoV/Japan/TY/WK-521/2020 from ToKyo (Japan) while the other 43 strains of SARS-CoV-2 had 29198C. Since RaTG13 and MP789 both had 8824C/28247T/29198T, we could speculate that these 8 strains of 29198T appeared earlier than strains of 29198C. Using this method, we reckoned that the earliest strains of SARS-CoV-2 was one patient admitted in Shenzhen hospital on 2020-1-10, whose entire type was 8824C/28247T/29198T/2682C/3812C/9606C/11125G/15667T/29808G. Besides, the earliest strain in the USA was from Phoenix, Arizona on 2020-1-26, whose entire type was 8824C/28247T/29198T/2682C/3812C/9606T/11125G/15667C/ 29808G. Patients with these two earliest strains both had exposure history to Wuhan. Therefore, we surmised that the origin of SARS-CoV-9 was still in Wuhan, but the earliest virus was still vague due to lack of enough samples (Supplementary Figure 1).

Furin is well recognized as an important serine protease, which has a minimum enzyme restriction site of Arg(R)-X-X-Arg (R) and plays an essential role in influenza infection. The binding capacity change of Furin in Avian Influenza may influence its pathogenicity^37^. Though Furin is not the most common protease in coronavirus, previous studies indicated its pivotal roles in SARS and MERS^28, 38^. RaTG13 was the strain closest to SARS-CoV-2 with 96% sequence similarity^39^. However, SRAS-CoV-2 had PRRA insertion in 690 amino acid site of S protein, with high conservation^40^. This insertion may become critical point for the host of RaTG13 turning from animal to human. Through sequence alignment, we found this inserted sequence may arise from the translocation between human coronavirus HKU1 and OC43 (Figure 4).

SARS-CoV-2 had three FCS (F1-3), where F1 hydrolyzes S protein to S1 and S2 and promotes virus-cell fusion, F2 hydrolyzes S2 and participates in virus pathogenicity after cell entry, while F3 functions through NTD and promotes adhesion between virus and cell surface. However, whether F3 site really exists and what is the function need further investigation. Furthermore, the target cell binding site of HKU1 and OC43 was on their S^A^ segment of S protein while its corresponding site in SARS-CoV-2 was NTD. Therefore, except for the potential interaction in F1 site, there also exists the possibility of interaction in NTD segment between SARS-CoV-2 and HKU1/OC43.

Viruses frequently undergo mutation and adjust its RSCU under evolutionary selection pressure to adapt to the host, facilitating better replication and dissemination^41^. The choice of Furin cleavage site might be an outstanding marker for coronavirus evolution. Meanwhile, we found that the mutation in ZJ01 may cause binding force change of FCS. All these data indicate that Furin might play a pivotal role in the pathogenicity of SARS-CoV-2. The evolution trends of increasing FCS in SARS-CoV-2 we found in this study are more prone to influenza-like clinical manifestations such as human HKU1 and OC43^42^.

Single cell sequencing analysis showed that Furin had higher expression level and wider organ distribution than ACE2, especially in salivary gland, lachrymal gland, colon, liver and kidney. Therefore, SARS-CoV-2 might evolve to utilize this specific feature by increasing FCS to become more infectious on multi-organ levels. Our hypothesis was in consistent with changed clinical characteristics of COVID-19 from published data and our observation, including detection of virus in feces^34^ and conjunctival secretions^35^, decreased severity/fatality, increased liver/kidney damage and symptoms of gastrointestinal tract, increased transmissibility and prolonged period of nuclear acid positivity. Since ACE2 expression was quite low in the whole body including lung, we speculated that, on one hand, it might be the inflammatory reaction whereas not the viral road itself triggering severe respiratory damage; on the other hand, the utilization of Furin may help scatter virus attack from lung to other organs, contributing to the phenomenon of decreased severity but increased liver/kidney dysfunction. Nevertheless, all these speculations warrant further investigations.

To sum up, ZJ01 isolated form mild COVID-19 patient of Zhejiang province represents a potential branch in virus evolution. SARS-CoV-2 may adopt a similar mechanism by depending on Furin for invasion as HJU1 and OC43. Such potential evolution direction may promote the appearance of mild subtype of COVID-19.

## Data Availability

Anyone can share this material, provided it remains unaltered in any way, this is not done for commercial purposes, and the original authors are credited and cited.

## Supplementary figure

**Supplementary figure 1.**
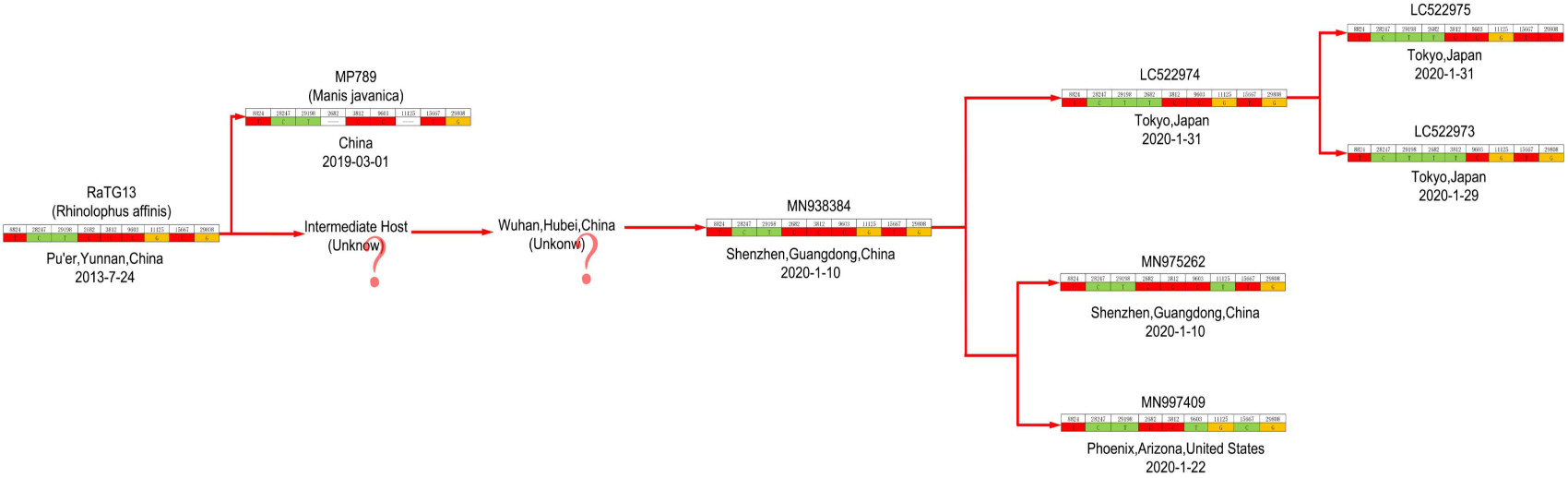
The evolution route of SARS-CoV-2 drawn based on T type tracing. RaTG13 was the most possible ancestor of SARS-CoV-2. In this study, the closest strain to RaTG13 was MN938384 that was isolated on 2020-1-10 from Guangdong province, China, which was more primitive than the T types from the USA and Japan. The patient with MN938384 had exposure history to Wuhan but there was no strain identified with such primitive feature in Wuhan. Therefore, we may confirm the origin of SARS-CoV-2 through enlarged and thorough screening the strains of T type in Wuhan.

## Funding

This research was supported by the National Key Program for Infectious Diseases of China (2017ZX10202202),the National Natural Science Foundation of China (No. 81770574, 81870080) and the National Key R&D Program of China, Stem Cell and Translation Research (2018YFA0109300).

## Competing interests

None declared.

## Acknowledgement

We thank Health Commission of Zhejiang Province, China for coordinating data collection; Thanks to all the front-line medical staffs of Zhejiang Province for their bravery and efforts in SARS-CoV-2 prevention and control.

## Notes

### Competing Interest Statement

The authors have declared no competing interest.

## Reference

1. Zhu N, Zhang D, Wang W, Li X, Yang B, Song J, et al. A Novel Coronavirus from Patients with Pneumonia in China, 2019. The New England journal of medicine 2020, 382(8): 727–733.

2. Gorbalenya AE BS, Baric RS, de Groot RJ, Drosten C, Gulyaeva AA, Haagmans BL, Lauber C, Leontovich AM, Neuman BW, Penzar D, Perlman S, Poon LL, Samborskiy D, Sidorov IA, Sola I, Ziebuhr J. Severe acute respiratory syndrome-related coronavirus: The species and its viruses-a statement of the Coronavirus Study Group. bioRxiv 2020.

3. Holshue ML, DeBolt C, Lindquist S, Lofy KH, Wiesman J, Bruce H, et al. First Case of 2019 Novel Coronavirus in the United States. The New England journal of medicine 2020.

4. Silverstein WK, Stroud L, Cleghorn GE, Leis JA. First imported case of 2019 novel coronavirus in Canada, presenting as mild pneumonia. Lancet (London, England) 2020.

5. Bogoch, II, Watts A, Thomas-Bachli A, Huber C, Kraemer MUG, Khan K. Potential for global spread of a novel coronavirus from China. Journal of travel medicine 2020.

6. Wu JT, Leung K, Leung GM. Nowcasting and forecasting the potential domestic and international spread of the 2019-nCoV outbreak originating in Wuhan, China: a modelling study. Lancet (London, England) 2020.

7. Kim JY, Choe PG. The First Case of 2019 Novel Coronavirus Pneumonia Imported into Korea from Wuhan, China: Implication for Infection Prevention and Control Measures. 2020, 35(5): e61.

8. Huang C, Wang Y, Li X, Ren L, Zhao J, Hu Y, et al. Clinical features of patients infected with 2019 novel coronavirus in Wuhan, China. Lancet (London, England) 2020.

9. Nanshan Chen MZ, Xuan Dong, Jieming Qu, Fengyun Gong, Yang Han, Yang Qiu, Jingli Wang, Ying Liu, Yuan Wei, Jia’an Xia, Ting Yu, Xinxin Zhang, Li Zhang. Epidemiological and clinical characteristics of 99 cases of 2019 novel coronavirus pneumonia in Wuhan, China: a descriptive study. Lancet (London, England) 2020, S0140-6736(20): 7.

10. Xu XW, Wu XX, Jiang XG, Xu KJ, Ying LJ, Ma CL, et al. Clinical findings in a group of patients infected with the 2019 novel coronavirus (SARS-Cov-2) outside of Wuhan, China: retrospective case series. 2020, 368: m606.

11. Wu Z, McGoogan JM. Characteristics of and Important Lessons From the Coronavirus Disease 2019 (COVID-19) Outbreak in China: Summary of a Report of 72314 Cases From the Chinese Center for Disease Control and Prevention. Jama 2020.

12. Li Q, Guan X, Wu P, Wang X, Zhou L, Tong Y, et al. Early Transmission Dynamics in Wuhan, China, of Novel Coronavirus-Infected Pneumonia. New England Journal of Medicine 2020.

13. Yang Yang QL, Mingjin Liu, Yixing Wang, Anran Zhang, Neda Jalali, Natalie Dean, Ira Longini, M. Elizabeth Halloran, Bo Xu, Xiaoai Zhang, Liping Wang, Wei Liu, Liqun Fang. epidemiological and clinical features of the 2019 novel coronavirus outbreak in China. medRxiv 2020.

14. Zou L, Ruan F, Huang M, Liang L, Huang H, Hong Z, et al. SARS-CoV-2 Viral Load in Upper Respiratory Specimens of Infected Patients. The New England journal of medicine 2020.

15. Walls AC, Xiong X, Park YJ, Tortorici MA, Snijder J, Quispe J, et al. Unexpected Receptor Functional Mimicry Elucidates Activation of Coronavirus Fusion. Cell 2019, 176(5): 1026-1039.e1015.

16. Hofmann H, Pohlmann S. Cellular entry of the SARS coronavirus. Trends in microbiology 2004, 12(10): 466–472.

17. WHO. Clinical management of severe acute respiratory infection when Novel coronavirus (nCoV) infection is suspected: interim guidance. Jan 11, 2020. https://wwwwhoint/internal-publications-detail/clinical-management-of-severe-acute-respiratory-infection-when-novel-coronavirus-(ncov)-infection-is-suspected (accessed Jan 30,2020) 2020.

18. Roujian Lu XZ, Juan Li, Peihua Niu, Bo Yang, Honglong Wu, Wenling Wang. Genomic characterisation and epidemiology of 2019 novel coronavirus: implications for virus origins and recptor binding. Lancet (London, England) 2020, S0140-6736(20): 7.

19. Duckert P, Brunak S, Blom N. Prediction of proprotein convertase cleavage sites. Protein engineering, design & selection: PEDS 2004, 17(1): 107–112.

20. Laureanti J, Brandi J, Offor E, Engel D, Rallo R, Ginovska B, et al. Visualizing biomolecular electrostatics in virtual reality with UnityMol-APBS. 2020, 29(1): 237–246.

21. Madissoon E, Wilbrey-Clark A, Miragaia RJ, Saeb-Parsy K, Mahbubani KT, Georgakopoulos N, et al. scRNA-seq assessment of the human lung, spleen, and esophagus tissue stability after cold preservation. 2019, 21(1): 1.

22. Parikh K, Antanaviciute A, Fawkner-Corbett D, Jagielowicz M, Aulicino A, Lagerholm C, et al. Colonic epithelial cell diversity in health and inflammatory bowel disease. Nature 2019, 567(7746): 49–55.

23. MacParland SA, Liu JC, Ma XZ, Innes BT. Single cell RNA sequencing of human liver reveals distinct intrahepatic macrophage populations. 2018, 9(1): 4383.

24. Stuart T, Butler A, Hoffman P, Hafemeister C, Papalexi E, Mauck WM, 3rd, et al. Comprehensive Integration of Single-Cell Data. Cell 2019, 177(7): 1888-1902.e1821.

25. Korsunsky I, Millard N, Fan J. Fast, sensitive and accurate integration of single-cell data with Harmony. 2019, 16(12): 1289–1296.

26. H W. ggplot2: Elegant Graphics for Data Analysis. Springer-Verlag New York 2016.

27. Guan WJ, Ni ZY, Hu Y, Liang WH, Ou CQ, He JX, et al. Clinical Characteristics of Coronavirus Disease 2019 in China. The New England journal of medicine 2020.

28. Simmons G, Bertram S, Glowacka I, Steffen I, Chaipan C, Agudelo J, et al. Different host cell proteases activate the SARS-coronavirus spike-protein for cell-cell and virus-cell fusion. Virology 2011, 413(2): 265–274.

29. de Greef JC, Slutter B, Anderson ME, Hamlyn R, O’Campo Landa R, McNutt EJ, et al. Protective role for the N-terminal domain of alpha-dystroglycan in Influenza A virus proliferation. Proceedings of the National Academy of Sciences of the United States of America 2019, 116(23): 11396–11401.

30. Meng T CH, Zhang H, Kang Z, Xu D, Gong H, Wang J, Li Z, Cui X, Xu H, Wei H, Pan X, Zhu R, Xiao J, Zhou W, Cheng L, Liu J. The insert sequence in SARS-CoV-2 enhances spike protein cleavage by TMPRSS. bioRxiv 2020.

31. Qi F QS, Zhang S, Zhang Z. Single cell RNA sequencing of 13 human tissues identify cell types and receptors of human coronaviruses. bioRxiv 2020.

32. Wang D, Hu B, Hu C, Zhu F, Liu X, Zhang J, et al. Clinical Characteristics of 138 Hospitalized Patients With 2019 Novel Coronavirus-Infected Pneumonia in Wuhan, China. Jama 2020.

33. Bai Y, Yao L, Wei T, Tian F, Jin DY, Chen L, et al. Presumed Asymptomatic Carrier Transmission of COVID-19. Jama 2020.

34. Pan Y, Zhang D, Yang P, Poon LLM, Wang Q. Viral load of SARS-CoV-2 in clinical samples. The Lancet Infectious diseases 2020.

35. Xia J, Tong J, Liu M, Shen Y, Guo D. Evaluation of coronavirus in tears and conjunctival secretions of patients with SARS-CoV-2 infection. 2020.

36. Xiaolu Tang CW, Xiang Li, Yuhe Song, Xinmin Yao, Xinkai Wu, Yuange Duan, Hong Zhang, Yirong Wang, Zhaohui Qian, Jie Cui, Jian Lu. On the origin and continuing evolution of SARS-CoV-2. National Science Review.

37. Tse LV, Hamilton AM, Friling T, Whittaker GR. A novel activation mechanism of avian influenza virus H9N2 by furin. Journal of virology 2014, 88(3): 1673–1683.

38. Millet JK, Whittaker GR. Host cell entry of Middle East respiratory syndrome coronavirus after two-step, furin-mediated activation of the spike protein. Proceedings of the National Academy of Sciences of the United States of America 2014, 111(42): 15214–15219.

39. Zhou P, Yang XL, Wang XG, Hu B, Zhang L, Zhang W, et al. A pneumonia outbreak associated with a new coronavirus of probable bat origin. Nature 2020.

40. Xin Li GD, Wei Zhang, Jinsong Shi, Jiayuan Chen, Shunmei Chen, Shan Gao, Jishou Ruan. A furin cleavage site was discovered in the S protein of the Wuhan 2019 novel coronavirus. chinaXiv 2020.

41. Li G, Zhang W, Wang R, Xing G, Wang S, Ji X, et al. Genetic Analysis and Evolutionary Changes of the Torque teno sus Virus. International journal of molecular sciences 2019, 20(12).

42. Friedman N, Alter H, Hindiyeh M, Mendelson E, Shemer Avni Y, Mandelboim M. Human Coronavirus Infections in Israel: Epidemiology, Clinical Symptoms and Summer Seasonality of HCoV-HKU1. Viruses 2018, 10(10).

